# Asymptomatic monkeypox virus infections among male sexual health clinic attendees in Belgium

**DOI:** 10.1101/2022.07.04.22277226

**Authors:** Irith De Baetselier, Christophe Van Dijck, Chris Kenyon, Jasmine Coppens, Dorien Van den Bossche, Hilde Smet, Laurens Liesenborghs, Fien Vanroye, Tessa de Block, Antonio Rezende, Eric Florence, Koen Vercauteren, Marjan Van Esbroeck, the Monkeypox study group

## Abstract

**Background:** Monkeypox is transmitted by close contact with symptomatic cases, and those infected are assumed to be uniformly symptomatic. Evidence of subclinical monkeypox infection is limited to a few immunological studies which found evidence of immunity against orthopoxviruses in asymptomatic individuals who were exposed to monkeypox cases. We aimed to assess whether asymptomatic infections occurred among individuals who underwent sexually transmitted infection (STI) screening in a large Belgian STI clinic around the start of the 2022 monkeypox epidemic in Belgium.

**Methods:** Anorectal and oropharyngeal swabs collected for gonorrhoea/chlamydia screening from May 1 until May 31, 2022 were retrospectively tested by a monkeypox-specific PCR. Cases with a positive PCR result were recalled to the clinic for case investigation, repeat testing and contact tracing.

**Findings:** In stored samples from 224 men, we identified three cases with a positive anorectal monkeypox PCR. All three men denied having had any symptoms in the weeks before and after the sample was taken. None of them reported exposure to a diagnosed monkeypox case, nor did any of their contacts develop clinical monkeypox. Follow-up samples were taken 21 to 37 days after the initial sample, by which time the monkeypox-specific PCR was negative, likely as a consequence of spontaneous clearance of the infection.

**Interpretation:** The existence of asymptomatic monkeypox infection indicates that the virus might be transmitted to close contacts in the absence of symptoms. Our findings suggest that identification and isolation of symptomatic individuals may not suffice to contain the outbreak.

**Funding:** Institutional funding

**Research in context:** *Evidence before this study:* Similar to smallpox, monkeypox is transmitted through close contact with symptomatic cases, and 100% of those infected are assumed to develop symptoms. These features imply that an outbreak in the general population tends towards extinction with relatively minor hygienic measures, as observed in several outbreaks in endemic regions. If, however, asymptomatic transmission occurs, the outbreak becomes much more difficult to contain. We searched PubMed and Google Scholar for evidence of asymptomatic human monkeypox, using the search terms “monkeypox” AND (“asymptomatic” OR “subclinical”), and included peer-reviewed reports published until June 17, 2022. We identified seven original reports in three different epidemiological settings which reported indirect, immunological evidence of asymptomatic monkeypox infection in a small number of people who were exposed to the virus. We did not find any study that provided direct evidence of the virus in asymptomatic individuals.

*Added value of this study:* By retrospectively screening clinical samples collected for sexually transmitted infection screening in our centre throughout May 2022 with a monkeypox-specific PCR, we found evidence of asymptomatic monkeypox virus infection in three individuals.

*Implications of all the available evidence:* The existence of asymptomatic monkeypox infection indicates that the virus may be transmitted in the absence of symptoms. This risk can be further quantified by studying viral dynamics in contacts of symptomatic and asymptomatic monkeypox cases. Our findings suggest that identification and isolation of symptomatic individuals may not suffice to contain the outbreak.

## Introduction

Monkeypox is a viral disease that is endemic in several African countries.^1^ Much of what is known about its natural history stems from outbreaks in the general population in those countries. Based on these data, monkeypox virus (MPXV) is thought to be transmitted through close contact with symptomatic cases.^2^ All those infected with MPXV are assumed to develop symptoms^3^ and the house hold secondary attack rate is approximately 10%.^2^ These features imply that, in the absence of repeated animal-to-human transmission, an outbreak in the general population tends towards extinction with relatively minor hygienic measures, as observed in several outbreaks in endemic regions.^1,4^

The current monkeypox outbreak in non-endemic countries differs from those described earlier, with respect to the affected population and the clinical case picture. Indeed, the current outbreak appears to primarily affect men who have sex with men (MSM).^5^ Many present with symptoms that are largely limited to the anogenital region, and some have only minimal symptoms.^6^ In addition, some studies found high viral loads in anogenital samples, saliva and semen.^7^ This led to the hypothesis that MPXV may be sexually transmitted.^7,8^ By June 17, more than 2500 cases have been confirmed in this outbreak which is far more than in previous outbreaks in the general population in endemic countries (https://ourworldindata.org/monkeypox). As such, researchers have raised a number of questions that could explain this extraordinary surge in cases.^9^ One of these questions is: could infection with MPXV be asymptomatic and could asymptomatic infections contribute to monkeypox spread?^9^ This is a crucial point since if asymptomatic infections could be transmitted onwards, this would mean that identification and isolation of symptomatic cases might not be sufficient to end the outbreak.

Here, we report three asymptomatic MPXV infections which were discovered by retrospectively screening clinical samples collected for anorectal and oropharyngeal gonorrhoea/chlamydia testing among men visiting the largest sexually transmitted infection (STI) clinic in Belgium, in May 2022.

## Methods

### Samples

Between May 1 and May 31, 2022, 237 men underwent testing for anorectal and/or oropharyngeal gonorrhoea/chlamydia as part of routine care at the HIV/STI clinic of the Institute of Tropical Medicine Antwerp, Belgium. The sample types tested were anorectal swabs, oropharyngeal swabs, or a combination of a patient’ s first-void urine, oropharyngeal swab, and anorectal swab (pooled sample, as described^10^). Samples were processed with the Abbott Real Time CT/NG assay which includes the Abbott m2000sp for DNA extractions (Abbott Molecular Des Plaines, Illinois, USA). The original swab samples and their remnant DNA extracts were frozen (−20°C) or refrigerated (2-8°C), respectively, until processing in the current study.

### Monkeypox-specific PCR

Step 1: Remnant DNA extracts were tested for the presence of MPXV using previously described primer sets targeting the MPXV-TNF receptor gene.^11^ The Applied Biosystems Quantstudio PCR system was used for PCR amplification.

Step 2: In case of a positive PCR result on a remnant DNA extract, confirmation of the result was sought on the original sample, as follows. Samples were thawed and spiked with Phocine herpesvirus 1 (PhHV-1) as process control. DNA extraction was done with Maxwell® Promega, using 300 µL sample input and 75 µL elution volume. DNA extracts were amplified using the same PCR primersets as mentioned above.

Step 3: As a means to verify specificity of the MPXV-PCR, PCR template sizes were analysed by TapeStation 4150 (Agilent) with the HS D1000 kit.

### Case management

Patients with a positive MPXV-PCR result were informed about their diagnosis and recalled to the clinic for additional case investigation and contact tracing in accordance with WHO guidance.^12^ At that time, follow-up samples were taken for MPXV PCR from the site where the virus was initially detected.

### Ethical considerations

The study protocol was approved by the Institutional Review Board of the Institute of Tropical Medicine (1600/22). Written informed consent is available from all asymptomatic cases.

## Results

A total of 225 remnant DNA extracts were available from 224 different men. These included 163 pooled samples, 60 anorectal swabs and two oropharyngeal swabs. Initial MPXV-PCR (step 1) was positive on four DNA extracts: three anorectal swabs, and one pooled sample (Table 1). MPXV-PCR after repeat DNA extraction of the original samples in step 2 confirmed those PCR results on all three anorectal swabs, and on the anorectal (but not oropharyngeal) swab that constituted the pooled sample. PCR template size analysis showed a +/-143 bp DNA fragment on gel electrophoresis for all four samples confirming specific amplification of the targeted MPXV genomic region.

**Table 1:**
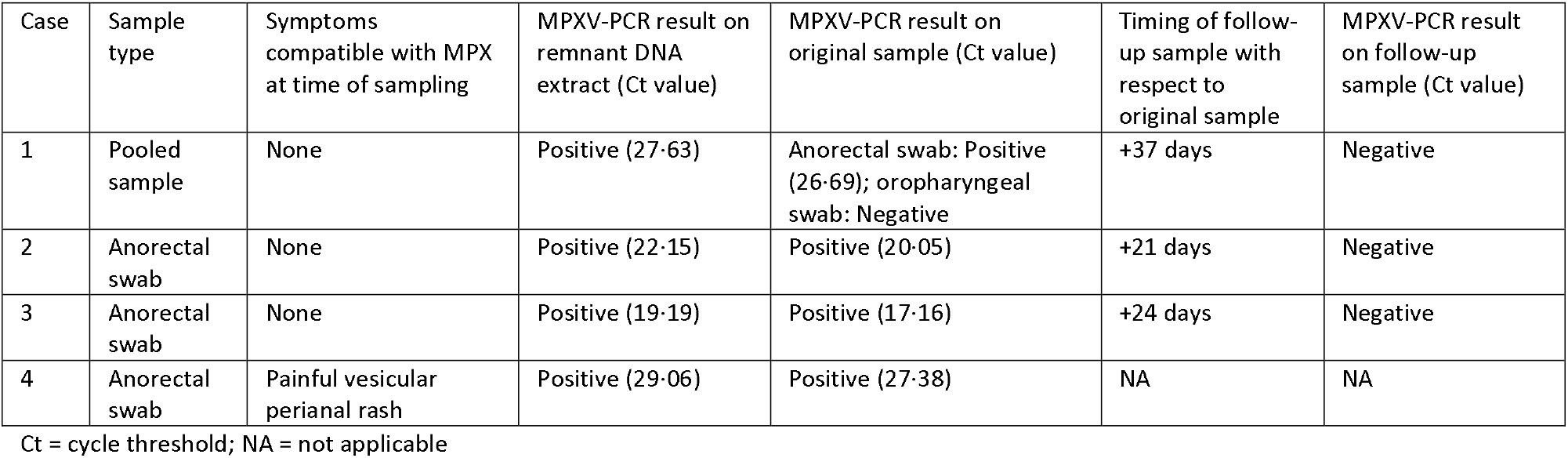
Characteristics of MPXV-positive samples

The four MPXV-positive samples in this study were collected from four men. At the time of sampling, one of those men suffered from a painful vesicular perianal rash, which was misdiagnosed as a flare-up of herpes simplex. The remaining three men did not report any symptoms at the time of sampling. According to the institutional guidelines for STI testing among asymptomatic individuals, no specific examination was done at the time of sampling and men self-collected their anorectal swabs at the clinic. Of note, all cases tested negative for N. gonorrhoeae/C. trachomatis.

The three asymptomatic monkeypox cases were men between 30 and 50 years old, each with a well-controlled HIV infection and a history of multiple STIs. None of them were vaccinated against smallpox. The three asymptomatic men were recalled to the clinic for further investigation within 21 to 37 days after the initial sampling. At that time, none of the men showed signs or symptoms of monkeypox and all denied having noticed any symptoms during the two months prior or 3 weeks after initial sampling. All three men had condomless sexual intercourse with at least one male partner within a few days to one month before sampling. Two out of three men had sexual contacts while travelling abroad within two weeks before sampling. As far as traceable, none of their partners have reported signs or symptoms of monkeypox. Results of basic laboratory investigations at the time of sampling, including renal and liver function tests, as well as C-reactive protein were normal. Follow-up anorectal swab samples tested PCR negative for MPXV (Table 1).

## Discussion

We found evidence of asymptomatic MPXV infection in three individuals, in the form of positive PCR results on anorectal samples. Even though the existence of asymptomatic MPXV infection has been suggested by a number of immunological studies in MPXV-exposed individuals,^13–19^ this was, to our knowledge, never substantiated by direct detection of the virus. Interestingly, one of the asymptomatic men in our study predates the first detected symptomatic case in Belgium by several days,^20^ and could not be epidemiologically linked to any other monkeypox case, nor did he report international travel or participation in mass gatherings. This may indicate that MPXV circulated among asymptomatic individuals in Belgium before the outbreak was detected.

Due to the retrospective nature of our study, we were unable to assess whether MPXV could also be detected in other body sites of asymptomatic individuals, such as the respiratory tract. MPXV-PCR cycle threshold values in anorectal samples of the asymptomatic men in this study were similar or lower than those in samples taken from typical monkeypox skin lesions that underwent the same testing procedure in our clinic (mean 24·0 +/-standard deviation (sd) 6·7, n = 52 samples, unpublished data). This indicates that the anorectal mucosa of asymptomatic cases may be as infectious as skin lesions of symptomatic cases. Similarly, anorectal cycle threshold values of symptomatic cases in our clinic were in the same range (mean 25·5 +/-sd 7·9, n = 43 samples, unpublished data), which presumably indicates similar viral loads. This would support the hypothesis that MPXV can be transmitted via anal sex, even in the absence of symptoms.

Asymptomatic carriership was previously thought to play a negligible role in the spread of orthopoxviruses. Despite the fact that smallpox virus could be detected in the upper respiratory tract of asymptomatic contacts of smallpox cases,^21^ smallpox eradication was primarily, and successfully, based on the identification and isolation of symptomatic cases.^22^ Also, during previous monkeypox outbreaks in endemic countries, the role of asymptomatic carriership was never demonstrated. However, it is believed that in monkeypox endemic settings, only a fraction of the true caseload is detected due to a lack of resources to do proper surveillance. Until now, there have been no systematic efforts to detect asymptomatic carriership of MPXV.

It is possible that in the current outbreak in non-endemic settings, asymptomatic carriership plays a more substantial role in virus transmission. In endemic settings, patients typically present with a more generalized rash and transmission occurs mainly within households or health care settings, presumably through direct contact with skin lesions or droplets.^2^ In the current outbreak, the skin eruption often remains localized at the site of inoculation, and the mode of transmission seems to be sexual.^8^ In this case, asymptomatic carriership, especially with high viral loads in the anal mucosa, could, therefore, be a significant driver of transmission.

The size of the current outbreak in non-endemic countries is larger than ever documented in the absence of repeated epizootic events. While this may be explained by the high number of close contacts among the affected cases,^23^ asymptomatic transmission to/from the anorectum in addition to other body sites may have played a role. Indeed, many reported cases so far had unprotected sex with one or several casual sex partners.^7,8,24,25^ Direct contact with breached anogenital mucosal membranes during sex may be a previously unrecognised mode of MPXV transmission.^26^ Viral transmission in the absence of noticeable symptoms could explain why self-isolation at symptom onset has been insufficient to halt the epidemic thus far.

We could not confirm MPXV-PCR positivity of the asymptomatic cases in a second sample taken from a different body site or at a different time point. By the time the MPXV-PCR result on the stored DNA extracts was available and the cases returned to the clinic to collect follow-up samples, the infection had likely been cleared. PCR specificity on the original results was supported by consistent PCR amplicon electrophoresis (supplementary figure 1). In addition, 98% of the MPXV genome was retrieved by whole genome sequencing (with an average 161.4x sequencing depth) in the original anorectal sample of case 2 (unpublished data). It should be noted that the three asymptomatic men in this study may not have been completely free of signs and symptoms of MPXV at the time of initial sampling, as no clinical or anoscopic examination found place at that time, and symptoms may not have been reported because of recall bias or because small and painless perianal lesions went unnoticed. These findings warrant thorough prospective serological, molecular and epidemiological studies involving monkeypox cases and their contacts. Such studies should elucidate the proportion of asymptomatic cases, to what extent asymptomatic cases are infectious, which body sites contribute to infectiousness, and whether condom use or vaccination could be used to prevent transmission from asymptomatic carriers.

These findings have important consequences for the management of the epidemic. Until the role of asymptomatic transmission is further elucidated, the precautionary principle applies.^23^ If we want to stop human-to-human transmission, control measures should be revisited. Firstly, awareness campaigns in the general and high risk populations should include the possibility of asymptomatic transmission among close (sexual) contacts.^23^ Secondly, efforts to identify asymptomatic cases should be increased, by contact tracing, and potentially by screening high risk populations. Finally, our data provide additional evidence to introduce vaccination of high risk populations.

## Data Availability

The data supporting the findings of this publication are retained at the Institute of Tropical Medicine (ITM), Antwerp and will not be made openly accessible due to ethical and privacy concerns. According to the ITM research data sharing policy, only fully anonymised data can be shared publicly. The data are de-identified (using a unique patient code) but not fully anonymised and it is not possible to fully anonymise them due to the longitudinal nature of the data. Data can however be made available after approval of a motivated and written request to the ITM at. The ITM data access committee will verify if the dataset is suitable for obtaining the study objective and assure that confidentiality and ethical requirements are in place.

## Authors’ contributions

IDB, MVE, KV, FV, DVDB conceptualized the study. HS performed the testing. IDB & CVD wrote the first draft of the manuscript with revision by KV & MVE. IDB & CVD analysed the data. CK recontacted the patients for more epidemiological information. All authors reviewed and approved the final version of the manuscript.

## Acknowledgements

We would like to thank the patients to provide additional information, and all colleagues of the clinical reference laboratory of the Institute of Tropical Medicine.

## References

1 Petersen E, Kantele A, Koopmans M, et al. Human Monkeypox: Epidemiologic and Clinical Characteristics, Diagnosis, and Prevention. Infect Dis Clin North Am 2019; 33: 1027–43.

2 Bunge EM, Hoet B, Chen L, et al. The changing epidemiology of human monkeypox—A potential threat? A systematic review. PLoS Negl Trop Dis 2022; 16: 1–20.

3 World Health Organization (WHO). Monkeypox Outbreak Toolbox: WHO suggested outbreak case definition. 2022. https://www.who.int/emergencies/outbreak-toolkit/disease-outbreak-toolboxes/monkeypox-outbreak-toolbox (accessed June 20, 2022).

4 Jezek Z, Grab B, Dixon H. Stochastic model for interhuman spread of monreypox. Am J Epidemiol 1987; 126: 1082–92.

5 Perez Duque M, Ribeiro S, Martins JV, et al. Ongoing monkeypox virus outbreak, Portugal, 29 April to 23 May 2022. Eurosurveillance 2022; 27: 1–6.

6 Ferraro F, Caraglia A, Rapiti A, et al. Letter to the editor : multiple introductions of MPX in Italy from different geographic areas. Eurosurveillance 2022; 27: 1–2.

7 Antinori A, Mazzotta V, Vita S, et al. Epidemiological, clinical and virological characteristics of four cases of monkeypox support transmission through sexual contact, Italy, May 2022. Euro Surveill 2022; 27: 1–6.

8 Heskin J, Belfield A, Milne C, et al. Transmission of monkeypox virus through sexual contact – A novel route of infection. J Infect 2022.

9 Dye C, Kraemer MUG. Investigating the monkeypox outbreak. BMJ 2022; 377: o1314.

10 De Baetselier I, Osbak KK, Smet H, Kenyon CR, Crucitti T. Take three, test one: a cross-sectional study to evaluate the molecular detection of Chlamydia trachomatis and Neisseria gonorrhoeae in pooled pharyngeal, anorectal and urine samples versus single-site testing among men who have sex with men in Belgium. Acta Clin Belgica Int J Clin Lab Med 2020; 75: 91–5.

11 Li Y, Zhao H, Wilkins K, Hughes C, Damon IK. Real-time PCR assays for the specific detection of monkeypox virus West African and Congo Basin strain DNA. J Virol Methods 2010; 169: 223–7.

12 World Health Organization (WHO). Surveillance, case investigation and contact tracing for Monkeypox. Interim guidance 22 May 2022..

13 Jezek Z, Marennikova SS, Mutumbo M, et al. Human monkeypox: A study of 2,510 contacts of 214 patients. J Infect Dis 1986; 154: 551–5.

14 Fenner F, Henderson DA, Arita I, Jezek Z, Ladnyi ID. Human Monkeypox and Other Poxvirus Infections of Man. In: Smallpox and its Eradication. 1988: 1287–319.

15 Jezek Z, Nakano JH, Arita I, Mutombo M, Szczeniowski M, Dunn C. Serological survey for human monkeypox infections in a selected population in Zaire. J Trop Med Hyg 1987; 90: 31–8.

16 Hammarlund E, Lewis MW, Carter S V., et al. Multiple diagnostic techniques identify previously vaccinated individuals with protective immunity against monkeypox. Nat Med 2005; 11: 1005–11.

17 Fleischauer AT, Kile JC, Davidson M, et al. Evaluation of human-to-human transmission of monkeypox from infected patients to health care workers. Clin Infect Dis 2005; 40: 689–94.

18 Karem KL, Reynolds M, Hughes C, et al. Monkeypox-induced immunity and failure of childhood smallpox vaccination to provide complete protection. Clin Vaccine Immunol 2007; 14: 1318–27.

19 Guagliardo SAJ, Monroe B, Moundjoa C, et al. Asymptomatic orthopoxvirus circulation in humans the wake of a monkeypox outbreak among chimpanzees in Cameroon. Am J Trop Med Hyg 2020; 102: 206–12.

20 Selhorst P, Rezende AM, de Block T, et al. Belgian case of Monkeypox virus linked to outbreak in Portugal. 2022. https://virological.org/t/belgian-case-of-monkeypox-virus-linked-to-outbreak-in-portugal/801 (accessed June 20, 2022).

21 Sarkar JK, Mitra AC, Mukherjee MK, D. SK. Virus excretion in smallpox. Bull World Health Organ 1973; 48: 523–7.

22 Fenner F. The global eradication of smallpox. Med J Aust 1980; 1: 455–6.

23 Pan D, Sze S, Nazareth J, et al. Monkeypox in the UK: arguments for a broader case definition. Lancet 2022; 6736: 1–2.

24 Hammerschlag Y, Macleod G, Papadakis G, et al. Monkeypox infection presenting as genital rash, Australia, May 2022. Eurosurveillance 2022; 27: 1–4.

25 de Nicolas-Ruanes B, Vivancos M, Azcarraga-Llobet C, et al. Monkeypox virus case with maculopapular exanthem and proctitis during the Spanish outbreak in 2022. J Eur Acad Dermatology Venereol 2022.

26 Miura F, Ewijk CE Van, Backer JA, Xiridou M, Franz E, Coul E Op De. Estimated incubation period for monkeypox cases confirmed in the Netherlands, May 2022. Eurosurveillance 2022; 27: 1–4.

