## supplementary figure 1 for "Asymptomatic monkeypox virus infections among male sexual health clinic attendees in Belgium"

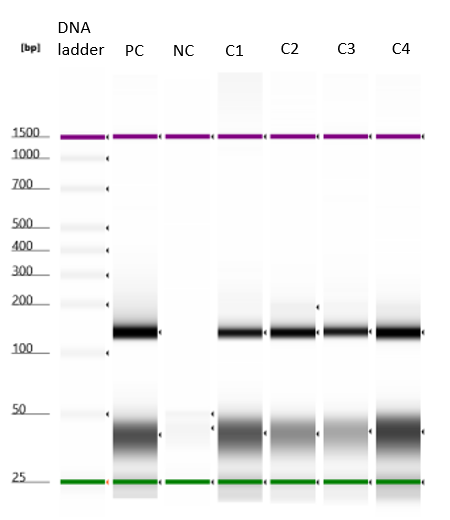

Supplementary Figure 1: PCR template sizes using Tapestation 4150 (Agilent). PC: Positive control; NC: Negative control; C: case
